# A Digital Diagnostic Pathway for Heart Failure: an economic evaluation

**DOI:** 10.1101/2024.11.01.24316568

**Authors:** Nicola Mcmeekin, Andrew Davies, Mark C Petrie, Ross T Campbell, David J Lowe, Clare L Murphy, Leeanne Macklin, Katriona Brooksbank, Olivia Wu

**Affiliations:** Health Economics and Health Technology Assessment (HEHTA), University of Glasgow; School of Cardiovascular and Metabolic Health, University of Glasgow; Queen Elizabeth University Hospital, NHS Greater Glasgow and Clyde; Forth Valley Royal Hospital, NHS Forth Valley; School of Health and Wellbeing, University of Glasgow

**Keywords:** economic evaluation, heart failure, diagnostic pathways, heart failure diagnosis, cost-consequence analysis, cost-effectiveness

## Abstract

Early diagnosis of heart failure improves patient outcomes, however diagnosis is complex and often delayed. This study evaluates a digital pathway for heart failure diagnosis, showing it reduces time to diagnosis and treatment initiation, with an incremental cost of £5,700 per quality adjusted life-year (QALY) gained over a lifetime. The pathway is cost-effective for the UK National Health Service at £20,000 per QALY threshold, however data uncertainties remain.

NPJ digital health 3000-4000 word limit plus abstract 70 words no sub-headings

## Introduction

Early diagnosis and treatment of heart failure (HF) can slow disease progression, avoid hospitalisations, and improve outcomes(1). However, diagnosing HF is complex and often delayed; as a result, prognosis for people with HF is poor. A recent meta-analysis reported survival rates at one, five and ten years to be 87%, 57% and 35%, respectively(2). European and UK guidelines recommend the use of an N-terminal pro b-type natriuretic peptide (NT-proBNP) blood test for patients with suspected HF in the community, to determine whether and how urgently patients should be referred to standard transthoracic echocardiogram (TTE) for confirmatory diagnosis(3). In practice, patients can endure repeated visits to primary care and hospitals before referral, an often complex and lengthy process.

Recent advances in digital health technologies have the potential to improve the HF diagnostic pathway, including digital platforms and artificial intelligence(4). The advantages of digital pathways include: redesigning and streamlining the diagnostic pathway (with added benefit of data collection for audit purposes); the ability for clinicians to remotely review patient results providing equity of care across geographical areas and for patients who struggle to access healthcare; improving patient outcomes by reducing time to diagnosis and treatment initiation; freeing TTE capacity by reducing the number of patients with suspected HF referred for TTE; early diagnosis of non-HF morbidities, and optimising treatment of symptoms.

Our study explored the potential value of introducing a digitised HF diagnostic pathway compared to a usual care pathway, from the perspectives of the UK NHS and patients.

## Methods

### Overview

We developed a decision analytic model to compare the introduction of a digital pathway for HF diagnosis with current practice in usual care. Patients enter the model upon referral onto the HF diagnostic pathway following a NT-proBNP test. We modelled costs and outcomes using a cost-consequence analysis (CCA) and a cost-utility analysis (CUA), over 12 months and lifetime horizons, respectively. Both the Medical Research Council (MRC) guidance on developing and evaluating complex interventions(5) and the National Institute for Health and Care Excellence (NICE) Evidence Standards Framework (ESF) for digital health technologies(6) recognise the relevance of CCA when evaluating complex, multi-component interventions and when other outcomes in addition to quality adjusted life years (QALYs) are of interest to decision-makers.

### Population

Patients referred from primary care to the HF diagnostic pathway, with symptoms suggestive of new HF and with an elevated NT-proBNP (≥400pg/ml).

### Comparator and Intervention

The comparator is current practice in usual care pathway, based on the NICE HF diagnosis and management guideline(7) and informed by healthcare professionals working in HF diagnosis. Routine referrals should be seen within six weeks (NT-proBNP <2000pg/ml) and urgent referrals within two weeks (NT-proBNP >2000pg/ml). In practice, however, these referrals may take substantially longer. The HF diagnostic pathway then begins with patients receiving electrocardiogram (ECG) and TTE performed by a cardiac physiologist.

The results of these investigations are reviewed by a cardiologist soon after. Patients diagnosed with HF with reduced ejection fraction (HFrEF) and HF with preserved ejection (HFpEF) are invited back to secondary care for consultation with a HF clinician to discuss their diagnosis and have treatment initiated. However, patients diagnosed with HFpEF may alternatively be referred back to their GP with management advice.

The digital pathway is an optimised pathway, which includes a digital platform and ‘one-stop’ service for diagnosis and treatment initiation. Patients with raised NT-proBNP results (>400pg/ml) are referred onto the digital pathway. Referral details including blood test results, presentation of symptoms, and patient medical history are transferred from general practice and presented on the digital platform. HF clinicians remotely review these details at a stage termed Active Clinical Referral Triage (ACRT) within days of referral. Suspected HF patients are invited to attend a one-stop diagnostic service, either as a ‘routine’ or ‘urgent’ referral. Patients not suspected to have HF exit the diagnostic pathway. Diagnosis for patients with suspected HF based on ACRT is then made at the one-stop service based on ECG performed by a healthcare support worker and TTE performed by a cardiac physiologist, as under the usual care pathway. A cardiology nurse specialist (CNS) reviews patient information and the TTE results on the digital platform, and logs a preliminary diagnosis of HFrEF, HFpEF or no HF. Patients are informed of their diagnosis by the CNS, who immediately initiates appropriate HF medication. Subsequently, a cardiologist remotely reviews the CNS preliminary diagnosis and submits an electronic comprehensive management plan to the patient’s GP.

The intervention is informed by a digital pathway implemented during the Optimising a Digital Diagnostic Pathway for Heart Failure in the Community (OPERA) study. OPERA was a prospective observational study of consecutive patients who have been referred onto a digital pathway for confirmation of HF diagnosis following NT-proBNP testing (1 December 2020 to 31 August 2021), across five outpatient sites in Glasgow, UK (Clinical trials NCT04724200).

### Model

A decision analytic model was developed to assess the cost-effectiveness of the digital pathway (Figure 1). A decision tree accounts for outcomes under both pathways over an initial period of 12 months. Patients then enter a simple long-term Markov process with risks for mortality and heart failure hospitalisation conditional on their 12-month HF status (HFrEF, HFpEF or no HF), over a time horizon of a further 24 annual model cycles (Figure 2). This represents a total time horizon of 25 years for patients, approximating lifetime in this population. The standard UK discount rate of 3.5% is applied to both costs and health outcomes(8). The model is probabilistic, with relevant parameters entered as appropriate probability distributions.

**Figure 1.**
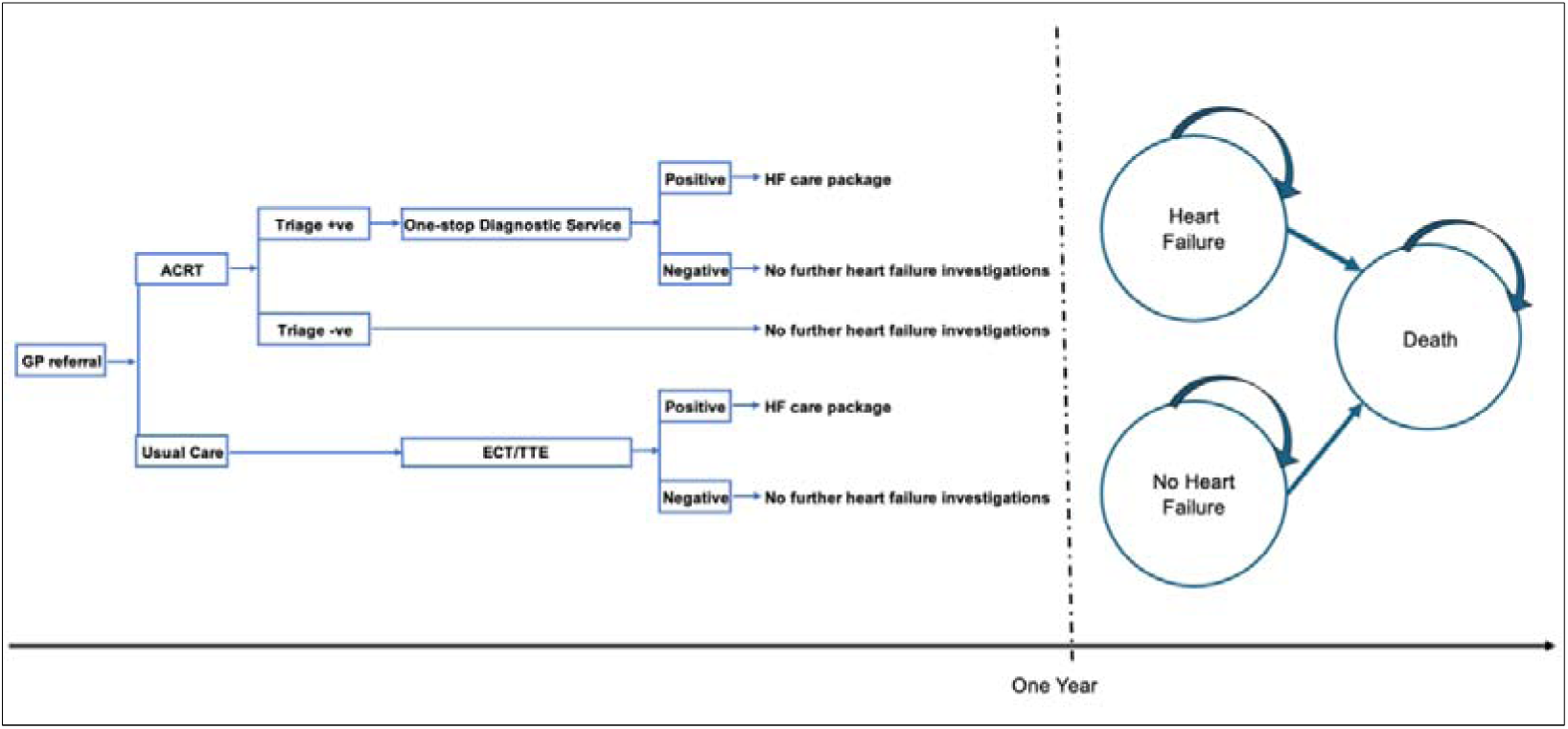
*Decision Analytic Model*

**Figure 2.**
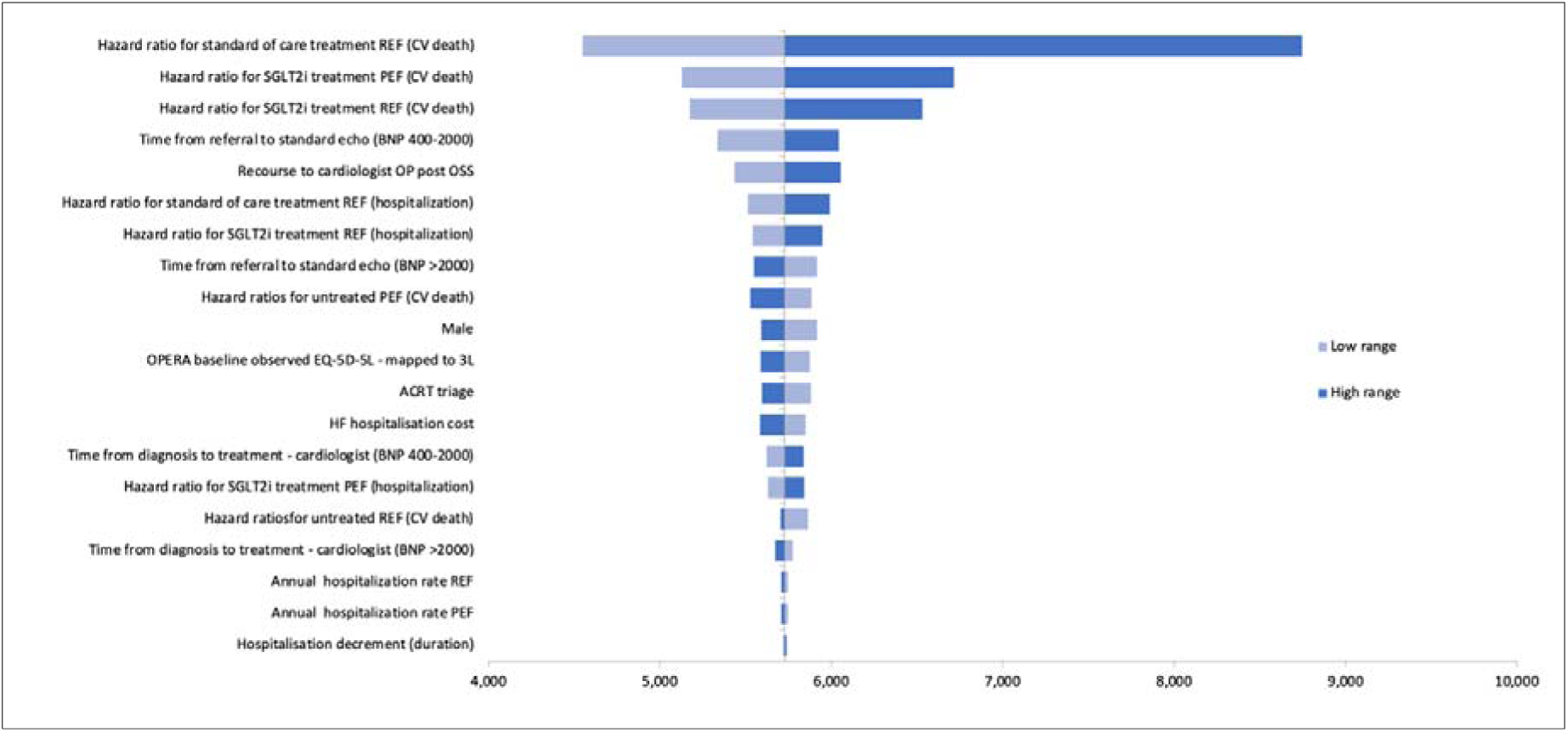
*Sensitivity Analyses*

### Clinical data and assumptions

Patients enter the model with baseline characteristics as observed in OPERA (Table 1), with mean age 75 years (SE 0.359) and 51% female. Most patients had NT-proBNP <2000pg/ml. The overall prevalence of HF was 81.5%, mainly with HFpEF, and NT-proBNP 400-2000pg/ml.

**Table 1.** Clinical and diagnostic model parameters

| Parameter |  | Value | SD or 95% Confidence Interval | Source |
| --- | --- | --- | --- | --- |
| **Patient Characteristics** |  |  |  |  |
|  | Age | 75 | N/A | OPERA |
|  | Female | 51% (n=439) | N/A |  |
| NT-proBNP 400-2000 (N, prevalence) | Not HF | 80 (18.4%) | N/A | OPERA |
|  | HFrEF | 66 (15.2%) | N/A |  |
|  | HFpEF | 219 (50.3%) | N/A |  |
| NT-proBNP >2000 (N, prevalence) | Not HF | 5 (1.1%) | N/A | OPERA |
|  | HFrEF | 24 (5.5%) | N/A |  |
|  | HFpEF | 41 (9.4%) | N/A |  |
| **Sensitivity \| Specificity** | TTE | 100% \| 100% | N/A | Assumption |
|  | ACRT | 100% \| 46% | N/A |  |
| **Timing of Pathway Elements** |  |  |  |  |
| Usual care | Referral for TTE | 180 days | ±27.0 | Assumption |
|  | Cardiologist review | 7 days |  | Assumption |
|  | Cardiologist OPC | 49 days |  | Assumption |
| Digital pathway | Referral to ACRT | 6 days | ±0.9 | Assumption |
|  | One-stop service | 50 days | ±7.5 | OPERA |
| **Heart Failure Hospitalisations** |  |  |  |  |
| Annual rate of admission | HFrEF | 0.219 | 0.202-0.237 | SOLVD |
|  | HFpEF | 0.044 | 0.040-0.049 | I_PRESERVE |
| HR hospitalisation HFrEF * | Standard of Care | 0.58 | 0.475-0.708 | EMPHASIS-HF |
|  | SGLT2i | 0.70 | 0.590-0.830 | DAPA-HF |
| HR hospitalisation HFpEF | SGLT2i | 0.71 | 0.600-0.830 | EMPEROR-PRESERVED |
| **Heart Failure Mortality** |  |  |  |  |
| HR Untreated HF vs population | HFrEF | 9.75 | 6.612-14.369 | SOLVD |
|  | HFpEF | 2.15 | 1.461-3.174 | I_PRESERVE |
| Treated HFrEF (versus untreated)* | Standard care | 0.44 | 0.278-0.696 | Burnett et al (2017) |
|  | SGLT2i | 0.82 | 0.690-0.980 | DAPA-HF |
| Treated HFpEF (versus untreated) | SGLT2i | 0.91 | 0.760-1.090 | EMPEROR-PRESERVED |
| Distributions: age/sex: none; NT-proBNP and prevalence: Dirichlet []; ACRT specificity: beta; pathway timings: gamma; hazard ratios: lognormal. |  |  |  |  |

We assumed that none of the patients with HF would be triaged out of the pathway at ACRT review stage. We assumed that ACRT would accurately triage out 30% of all patients, all of whom without HF(9). The total number of patients who entered ACRT was not recorded in OPERA; 825 patients remained in the diagnostic pathway following ACRT (i.e. assumed 70% of all patients who entered ACRT), of whom 421 were ultimately confirmed not to have HF. This is equivalent to the assumption that overall, 46% of all patients without HF who entered ACRT would be correctly diagnosed with no HF.

Empirical data on time to each point in the diagnostic pathways is not available. The model relies on estimates provided by cardiologists and CNS involved in the OPERA study. In the usual care pathway, the time from referral to TTE is estimated to be 180 days, with a further 56 days for cardiologist review and subsequent cardiologist consultation. Under the digital pathway ACRT is estimated to take place within six days. The interval from ACRT to the one-stop diagnostic and treatment initiation service of 50 days is based on data collected in OPERA. Given the uncertainties regarding the accuracy of ACRT in identifying non-HF patients and in usual care timelines, both elements are a key focus in our analyses.

We based estimates of untreated HF hospitalisation rates on SOLVD for HFrEF(10) and I-PRESERVE for HFpEF(11). Reduction in these risks was attributed to treatment with standard of care triple therapy for HFrEF based on EMPHASIS-HF(12), with further benefit from treatment with SGLT2i for HFrEF based on DAPA-HF(13), and SGLT2i based on the EMPEROR preserved trial(14). HF medication confers risk reductions for HF hospitalisation from initiation of treatment.

Risk of mortality is applied to all patients based on national life tables for Scotland(15), with HF patients modelled to be at elevated risk based on hazard ratios in comparison to the general population depending on ejection fraction, as in SOLVD and I-PRESERVE(10, 11). Once treatment is initiated, however, risk reductions are applied, again based on EMPHASIS-HF, DAPA-HF, and EMPEROR preserved. Some patients newly diagnosed with HF may already be treated with HF relevant medication. However, we assume that this will not be optimised for HF (the diagnosis not having previously been made). We assume such prior medication would have negligible effect on HF mortality.

When using control group data for untreated hospitalisation and mortality from SOLVD(10)and I-PRESERVE(11) we considered the following: 7% of the participants in the SOLVD trial were taking beta blockers at baseline so the effects were likely small from these, and whilst 85% were taking diuretics, the effect of these on lowering mortality are likely to be small. Also, diuretics are not one of the current usual therapy medications for heart failure. The baseline medications the participants in the I-PRESERVE trial were receiving would be for co-morbidities and would not impact heart failure prognosis.

### Health related quality of life

Mean health utilities measured by EQ-5D-3L in OPERA for those ultimately diagnosed with HF was 0.620 (SE 0.014), representing a 17% decrement relative to the UK population norm (16) of 0.755. For patients without HF we applied a UK population norm accounting for reductions with increasing age and maintained the same relative reduction for those with HF throughout the model. Though HF medications have been shown in clinical trials to improve quality of life as measured for example by the Kansas City Cardiomyopathy Questionnaire (KCCQ), we assumed no benefit of HF medication in terms of health-related quality of life. We did, however, apply decrements to HF hospitalisations based on analysis of the SHIFT study(17). That analysis controlled for SHIFT patients’ NYHA grade and estimated approximately a 0.23 (SE 0.155) reduction in quality of life for HF hospitalisation; whereas the analysis applied a decrement over a period 30 days prior to and 30 days after a hospitalisation, we assume a simple 30 days in total for each hospitalisation.

### Resource use

Resource use for each patient referred onto the usual care pathway comprises an ECG and a TTE, an associated cardiologist review (assumed to be ten minutes per review), and a cardiologist out-patient consultation for final diagnosis and treatment initiation. For the digital pathway, we assumed ten minutes cardiologist time per ACRT review, an ECG and a TTE at the one-stop diagnosis service, with 15 minutes of a healthcare support worker’s time, 45 minutes of a cardiac physiologist’s time, 45 minutes of an CNS’s time, and 15 minutes cardiologist time to remotely review each patient’s results. We also assumed that 25% of patients attending the one-stop service will subsequently require a follow-up consultation with a cardiologist.

Unit costs are presented in Table 2. Costs for the digital platform (Lenus Health) include capital costs (integration of IT equipment in pathway), an annual license (which covers 750 patients using the one-stop diagnostic service) and additional user fees (when the number of patients exceeds 750). In the model a unit cost per patient is assigned assuming 1,500 patients are referred to the heart diagnosis pathway annually. Other resource use through the pathways were based on standard UK unit cost sources; 2021 costs (£GBP) were applied to resource estimates to calculate the average cost for each pathway. Medication costs are based on British National Formulary (22). In the case of HFrEF and HFpEF the standard of care four pillar treatment regimen (beta-blockers, aldosterone receptor antagonists valsartan, SGLT2 inhibitors), or SGLT2 inhibitor alone is assumed respectively.

**Table 2.**
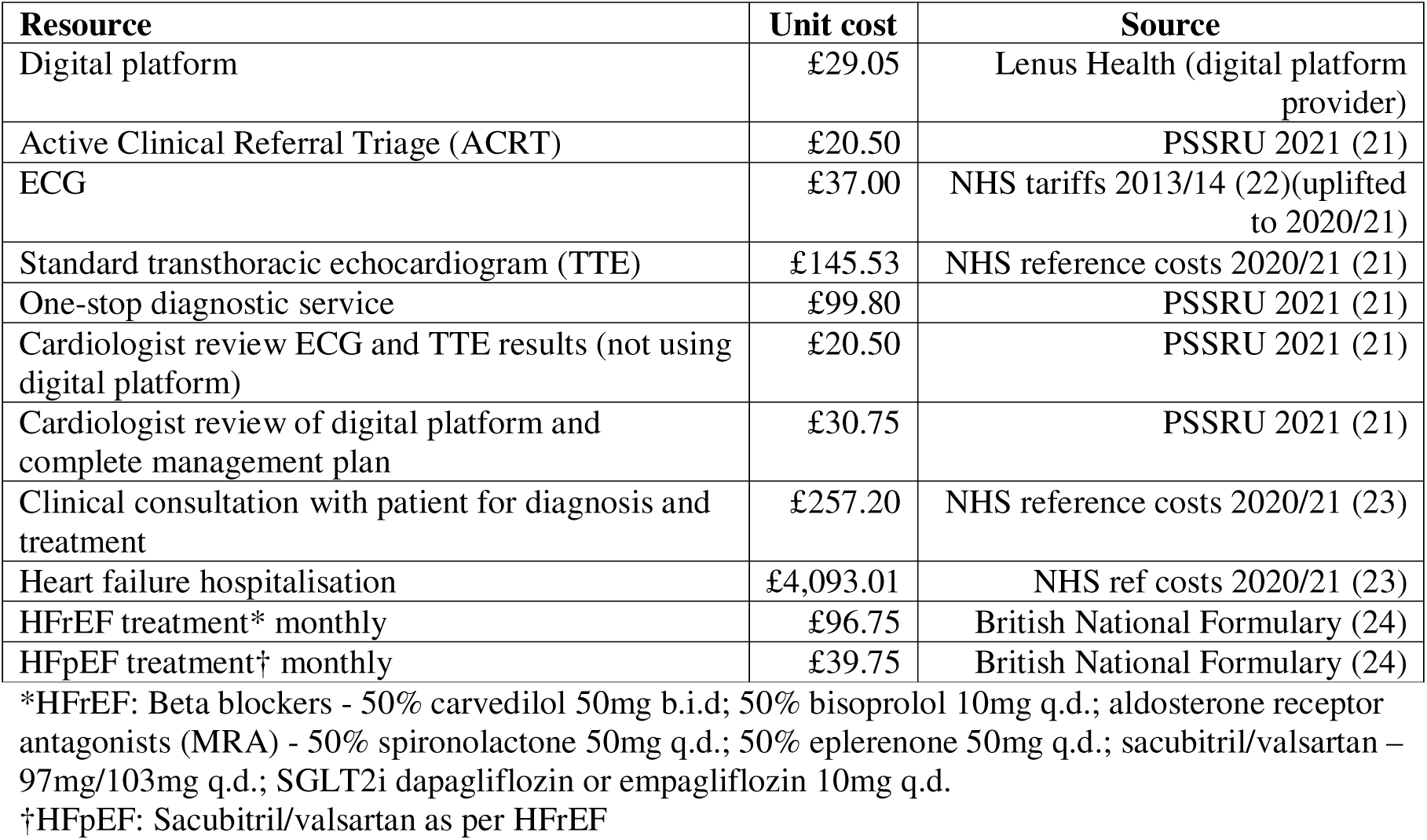
Unit costs

| **Resource** | **Unit cost** | **Source** |
| --- | --- | --- |
| Digital platform | £29.05 | Lenus Health (digital platform provider) |
| Active Clinical Referral Triage (ACRT) | £20.50 | PSSRU 2021 (21) |
| ECG | £37.00 | NHS tariffs 2013/14 (22)(uplifted to 2020/21) |
| Standard transthoracic echocardiogram (TTE) | £145.53 | NHS reference costs 2020/21 (21) |
| One-stop diagnostic service | £99.80 | PSSRU 2021 (21) |
| Cardiologist review ECG and TTE results (not using digital platform) | £20.50 | PSSRU 2021 (21) |
| Cardiologist review of digital platform and complete management plan | £30.75 | PSSRU 2021 (21) |
| Clinical consultation with patient for diagnosis and treatment | £257.20 | NHS reference costs 2020/21 (23) |
| Heart failure hospitalisation | £4,093.01 | NHS ref costs 2020/21 (23) |
| HFrEF treatment* monthly | £96.75 | British National Formulary (24) |
| HFpEF treatment† monthly | £39.75 | British National Formulary (24) |

\*HFrEF: Beta blockers - 50% carvedilol 50mg b.i.d; 50% bisoprolol 10mg q.d.; aldosterone receptor antagonists (MRA) - 50% spironolactone 50mg q.d.; 50% eplerenone 50mg q.d.; sacubitril/valsartan – 97mg/103mg q.d.; SGLT2i dapagliflozin or empagliflozin 10mg q.d.
†HFpEF: Sacubitril/valsartan as per HFrEF

### Analysis

We modelled short term (within 12 months) resource use and survival, and lifetime cost per QALY. The main analyses were performed probabilistically. Sensitivity analyses are presented based on the deterministic model.

## Results

Results for short- and long-term are presented in Table 3.

**Table 3.** Outcomes and Cost-effectiveness

|  | **Digital pathway** |  | **Usual care pathway** |  |
| --- | --- | --- | --- | --- |
| **One-year outcomes** |  |  |  |  |
|  | **Mean Units** | **£** | **Mean Units** | **£** |
| ACRT | - | - | 0.998 | 80 |
| Echo | 0.943 | 192 | 0.895 | 163 |
| One-stop service | - | - | 0.895 | 89 |
| Clinic visit (HF clinician) | 0.787 | 202 | 0.201 | 52 |
| Sub-total |  | 394 |  | 385 |
| Hospitalisation | 0.055 | 225 | 0.042 | 170 |
| Time on medication |  | 191 |  | 456 |
| Total one-year cost |  | 810 (651 - 969) |  | 1,011 (917 – 1,116) |
| Time to diagnosis (weeks) | 32 (26 - 40) |  | 7 (6 - 9) |  |
| One-year mortality (%) | 0.095 (0.073 - 0.12) |  | 0.076 (0.057 - 0.099) |  |
| **Long-term** |  |  |  |  |
| Diagnosis (year 1) |  | 394 (380 - 407) |  | 385 (377 - 392) |
| Hospitalisation |  | 839 (529 – 1,229) |  | 815 (498 – 1,226) |
| Medication |  | 2,073 (1,342 – 2,893) |  | 2,430 (1,683 – 3,326) |
| Total costs (£) |  | 3,306 (2,316 – 4,445) |  | 3,629 (2,629 – 4,837) |
| Life years |  | 6.409 (5.033 - 8.072) |  | 6.514 (5.093 - 8.246) |
| QALYs |  | 3.588 (2.880 - 4.369) |  | 3.645 (2.910 - 4.447) |
| Cost-per QALY (£) |  |  |  | 5,732 (4,351 – 9,650) |

### Short Term Cost-consequence Analysis

Over one-year, 87.4% of patients entering the digital pathway were expected to attend the one stop service and undergo echocardiography, compared to an expected 94.4% of patients in the usual care pathway undergoing echocardiography, with 78.8% of patients in the usual care pathway going on to consultation with a HF clinician. Total one year diagnosis costs are modelled to be £67 lower for the digital pathway.

For those ultimately diagnosed with HF time to treatment and the time to diagnosis was shorter with the digital pathway and led to earlier initiation of HF treatment at a mean of 50.4 days for the digital pathway compared with 225 days in the usual care pathway. Earlier confirmation of appropriate treatment resulted in higher medication costs for the first year in the digital pathway of £455 compared to £190 for usual care. Earlier treatment resulted in fewer hospitalisations and lower first year mortality of 7.4% for the digital pathway compared to 9.4% for usual care. Total first year costs, including hospitalisations, were £142 higher for the digital pathway.

### Cost-utility Analysis

Reduced one year mortality in the digital pathway was extrapolated to lead to a mean increase in (discounted) life expectancy of 0.105 years, or 0.056 QALYs. Greater one year survival also resulted in higher lifetime treatment costs for the digital pathway of approximately £350 per patient. The total incremental lifetime cost was modelled to be £323, with a cost per QALY for the digital pathway of £5,732 (95% confidence interval £4,351-£9,650), with a 100% probability of being cost-effective at a cost-effectiveness threshold of £20,000 per QALY.

Sensitivity analysis showed the digital pathway’s cost-effectiveness to be relatively insensitive to replacement of base case parameter values with their 95% confidence ranges (Figure 2). The digital pathway’s cost-effectiveness was most sensitive to the degree of benefit assigned for HF treatments, with the ICER reaching £8,748 when the upper 95% confidence interval for standard of care HR for cardiovascular death in HFrEF was applied. When the time from referral to standard echo in the usual care pathway was assumed to be 131 rather 180 days the digital pathway’s ICER fell marginally, to £5,333, as standard care medication costs rose as a result. We applied an estimate for the rate of triage from the digital pathway at ACRT of 30; alternative estimates of 20% to 40% had negligible impact on the cost per QALY.

## Discussion

There is growing interest in the adoption of digital health technologies, to tackle increasing pressures on healthcare systems. Healthcare commissioners are faced with early adoption decisions based on limited available evidence. The introduction digital health technologies to the HF diagnostic pathway is one such example.

We have conducted a comprehensive economic evaluation of the digital HF pathway for diagnosis and treatment initiation compared to usual care. In the absence of empirical data on comparative effectiveness, we used best available evidence from literature and from the recent OPERA study. The OPERA study was a prospective observational cohort study where all patients with suspected HF presenting at primary care were referred to ACRT. The study has been reported to have reduced referrals for heart failure diagnostic tests, including echocardiography, allowed consultants to deliver 10 clinical management plans in the time required for four face to face consultations, and reduced heart failure diagnosis waiting times from 12 months to six weeks(9). We applied a mid-way point of 180 days for diagnosis waiting time under usual care. Nevertheless, this analysis illustrates the benefits both for healthcare systems and patients that may be achieved with adoption of a digital pathway. We presented the short-term costs and consequences under the alternative pathways in the manner of CCA, as advocated in complex interventions and digital intervention evaluation guidelines. Quantifying specific short-term outcomes such as consultant time savings and diagnosis waiting times may assist health services commissioners considering the implementation of innovations such as the digital pathway. Though both over the short-term, and lifetime analyses, the digital pathway was associated with additional total health care costs, this is due to the earlier prescription of appropriate medication, and the greater short-term survival as a consequence. Short-term survival is higher in the digital pathway due to earlier diagnosis, however over a life-time this incremental benefit reduces, because after one year we assume equal treatment benefits in both pathways. When considering only the costs for diagnosis, the digital pathway itself appears cost-saving.

There are several limitations to our study, primarily associated with the paucity of data in this context. Despite existing guidelines on the usual care pathway for HF diagnosis, due to healthcare systems issues, this is not implemented in practice. In addition, there is heterogeneity in HF diagnostic pathways within and between different regions, making identifying the usual care pathway difficult. We had to make assumptions on times from referral to diagnosis and to treatment within the usual care pathway that is reflective of current practice. We also made assumptions on the accuracy of TTE plus expert diagnosis (either cardiologist or advance nurse practitioner) and that TTE is the gold standard for diagnosis; however this is broadly in line with previous published studies (18, 19). We were unable to identify recent data on comparative untreated HF treatments for mortality and hospitalisations, and the data we identified for the model were old and may not be representative of current observations (SOLVD and I-PRESERVE). Whilst the authors acknowledge that some patients were taking medication in these trials, they concluded this would not impact on heart failure prognosis, would have minimal impact on mortality and was the best evidence available; participants in more recent trials, in both control and treatment arms, are receiving standard care for heart failure. This justification is in line with the approach used in the economic evaluation model informing current NICE heart failure management recommendations(19). Finally, we used general population health utilities in our model, this results in the health utilities of the HF negative group potentially being overestimated as the self-reported health utilities of this cohort may not be as good as the general population. We have conducted extensive sensitivity analysis to test our assumptions; results remained consistent with the base case analysis.

We have adopted conservative estimates on potential benefits of the digital HF pathway, where appropriate. There may be additional benefits associated with the digital platform that our analysis does not consider. In our analysis we assign equivalent risks for hospitalisation and mortality to all treated HF patients. However, digital pathway extends to virtual clinical management which can involve ongoing collection of data and optimisation of medication based on changing clinical characteristics. This may help avoid hospital admissions through earlier detection of worsening heart failure, preserve health related quality of life, and potentially reduce mortality.

There is potential to include artificial intelligence (AI) in the digital pathway to further improve efficiency of diagnosis(20). To facilitate this requires identifying where in the pathway to place AI, training on its use, and additional costs of providing this component of the pathway. Introducing AI could reduce the number of people referred onto the diagnostic pathway and further reduce those who go on to the one-stop service and use related healthcare resources. Whilst the possible service efficiencies are clear, patient outcomes would depend on the sensitivity and specificity of the AI component used.

## Conclusions

This economic evaluation has estimated of the value of a digital pathway, indicating adoption of an OPERA-style digital pathway is a cost-effective strategy for the NHS and the patients in Scotland. However, there are varying levels of uncertainties associated with limitations of available data. In order to mitigate risk of investment, decision makers might consider a coverage with evidence approach, where the health technology is adopted for a fixed time period with the requirement for additional data collection and an evaluation in the future.

## Data Availability

Study data and model will be shared on a reasonable request to the corresponding author.

## Acknowledgements

The authors of this paper would like to thank the following people for their support and input to the research: Malbinder Fagura (AstraZeneca plc) and Paul McGuinness (Lenus Health Ltd).

## Ethics approval

The OPERA study was performed according to the UK Policy Framework for Health and Social Care Research and the Declaration of Helsinki and was approved by the West of Scotland Research Ethics Committee (REC) (20/SW/0182) and the Health Research Authority (HRA).

## References

1. Skaner Y, Bring J, Ullman B, Strender L-E. Heart failure diagnosis in primary health care: clinical characteristics of problematic patients. A clinical judgement analysis study. BMC family practice. 2003;4:12-.

2. Jones NR, Roalfe AK, Adoki I, Hobbs FDR, Taylor CJ. Survival of patients with chronic heart failure in the community: a systematic review and meta-analysis. European Journal of Heart Failure. 2019;21(11):1306–25.

3. Cowie MR. The heart failure epidemic: a UK perspective. Echo Research and Practice. 2017;4(1):R15–R20.

4. Li X-M, Gao X-Y, Tse G, Hong S-D, Chen K-Y, Li G-P, et al. Electrocardiogram-based artificial intelligence for the diagnosis of heart failure: a systematic review and meta-analysis. Journal of Geriatric Cardiology. 2022;19(12):970–80.

5. Skivington K, Matthews L, Simpson SA, Craig P, Baird J, Blazeby JM, et al. Framework for the development and evaluation of complex interventions: gap analysis, workshop and consultation-informed update. 2021;25:57.

6. National Institute for Health and Care Excellence. Evidence Standards Framework for Digital Health Technologies 2019 [Available from: https://www.nice.org.uk/Media/Default/About/what-we-do/our-programmes/evidence-standards-framework/digital-evidence-standards-framework.pdf.

7. National Institute for Health and Care Excellence. Chronic heart failure in adults: diagnosis and management - NICE guideline [NG106] 2018 [Available from: https://www.nice.org.uk/guidance/ng106.

8. National Institute for Health and Care Excellence N. NICE health technology evaluations: the manual 2022 [Available from: https://www.nice.org.uk/process/pmg36/chapter/economic-evaluation.

9. NHS England. Transforming heart failure diagnosis pathway to improve the patient journey: @NHStransform; 2024 [Available from: https://transform.england.nhs.uk/key-tools-and-info/digital-playbooks/cardiology-digital-playbook/transforming-heart-failure-diagnosis-pathway-to-improve-the-patient-journey/.

10. Yusuf S. Effect Of Enalapril On Survival In Patients With Reduced Left-Ventricular Ejection Fractions And Congestive-Heart-Failure. New England Journal of Medicine. 1991;325(5):293–302.

11. Massie BM, Carson PE, McMurray JJ, Komajda M, McKelvie R, Zile MR, et al. Irbesartan in Patients with Heart Failure and Preserved Ejection Fraction. New England Journal of Medicine. 2008;359(23):2456–67.

12. Zannad F, McMurray JJV, Krum H, van Veldhuisen DJ, Swedberg K, Shi H, et al. Eplerenone in Patients with Systolic Heart Failure and Mild Symptoms. New England Journal of Medicine. 2011;364(1):11–21.

13. McMurray JJV, Solomon SD, Inzucchi SE, Kober L, Kosiborod MN, Martinez FA, et al. Dapagliflozin in Patients with Heart Failure and Reduced Ejection Fraction. New England Journal of Medicine. 2019;381(21):1995–2008.

14. Anker SD, Butler J, Filippatos G, Ferreira JP, Bocchi E, Boehm M, et al. Empagliflozin in Heart Failure with a Preserved Ejection Fraction. New England Journal of Medicine. 2021;385(16):1451–61.

15. National Records of Scotland. Life Expectancy in Scotland, 2018-2020 2020 [Available from: https://www.nrscotland.gov.uk/statistics-and-data/statistics/statistics-by-theme/life-expectancy/life-expectancy-at-scotland-level.

16. Ara R, Brazier JE. Using Health State Utility Values from the General Population to Approximate Baselines in Decision Analytic Models when Condition-Specific Data are Not Available. Value in Health. 2011;14(4):539–45.

17. Griffiths A, Paracha N, Davies A, Branscombe N, Cowie MR, Sculpher M. Analyzing Health-Related Quality of Life Data to Estimate Parameters for Cost-Effectiveness Models: An Example Using Longitudinal EQ-5D Data from the SHIFT Randomized Controlled Trial. Advances in Therapy. 2017;34(3):753–64.

18. Taylor C, Monahan M, Roalfe A, Barton P, Iles R, Hobbs FobotoR, et al. The REFER (REFer for EchocaRdiogram) study: a prospective validation and health economic analysis of a clinical decision rule, NT-proBNP or their combination in the diagnosis of heart failure in primary care. Efficacy Mech Eval. 2017;4(3).

19. National Institute for Health and Care Excellence. Chronic heart failure in adults: diagnosis and management - NICE guideline [NG106]: addendum-appendicies 2018 [Available from: https://www.nice.org.uk/guidance/ng106/documents/addendum-appendicies.

20. Yasmin F, Shah SMI, Naeem A, Shujauddin SM, Jabeen A, Kazmi S, et al. Artificial intelligence in the diagnosis and detection of heart failure: the past, present, and future. Reviews in Cardiovascular Medicine. 2021;22(4):1095–113.

21. Jones K, Burns A. Unit Costs of Health and Social Care 2021 PSSRU, University of Kent, Canterbury 2021 [Available from: https://www.pssru.ac.uk/project-pages/unit-costs/unit-costs-of-health-and-social-care-2021/.

22. UK Government. Payment by Results in the NHS: tariff for 2013 to 2014 2013 [Available from: https://www.gov.uk/government/publications/payment-by-results-pbr-operational-guidance-and-tariffs.

23. National Health Service (England). National Schedule of NHS Costs 2020/21 2021 [Available from: https://www.england.nhs.uk/publication/2020-21-national-cost-collection-data-publication/.

24. British National Formulary. BNF Publications 2023 [Available from: https://bnf.nice.org.uk/drugs/.

